# Recording of intellectual disability in general hospitals 2006-2019: cohort study using linked datasets

**DOI:** 10.1101/2022.09.30.22280555

**Authors:** Rory Sheehan, Hassan Mansour, Matthew Broadbent, Angela Hassiotis, Christoph Mueller, Robert Stewart, Andre Strydom, Andrew Sommerlad

**Affiliations:** Institute of Psychiatry, Psychology and Neuroscience, King’s College London, London, UK; Division of Psychiatry, University College London, London, UK

## Abstract

**Background:** Accurate recognition and recording of intellectual disability in those who are admitted to general hospitals is necessary for making reasonable adjustments, ensuring equitable access, and monitoring quality of care. In this study we determined the rate of recording of intellectual disability in those with the condition who were admitted to hospital, and factors associated with the condition being unrecorded.

**Methods and Findings:** Retrospective cohort study using two linked datasets of routinely collected clinical data. We identified adults with diagnosed intellectual disability in a large secondary mental healthcare database and used general hospital records to investigate recording of intellectual disability when people were admitted to general hospitals between 2006 and 2019. Trends over time and factors associated with intellectual disability being unrecorded were investigated. We obtained data on 2,477 adults with intellectual disability who were admitted to a general hospital in England at least once during the study period (total number of admissions=27,314; median number of admissions=5). People with intellectual disability were accurately recorded as having the condition during 2.9% (95%CI 2.7-3.1%) of their admissions. Broadening the criteria to include a non-specific code of learning difficulty increased recording to 27.7% (95%CI 27.2-28.3%) of all admissions. Having a mild intellectual disability and being married were associated with increased odds of the intellectual disability being unrecorded in hospital records. We had no measure of quality of hospital care received and could not relate this to the presence or absence of a record of intellectual disability in the patient record.

**Conclusions:** Recognition and recording of intellectual disability in adults admitted to English general hospitals needs to be improved. Staff awareness training, screening at the point of admission, and data sharing between health and social care services could improve care for people with intellectual disability.

## Introduction

Intellectual disability is a lifelong disorder characterised by deficits in general cognitive ability and impaired functional skills [1]. People with intellectual disability constitute between 1 and 2% of the population, equating to approximately one million people in England alone [2]. Adults with intellectual disability have worse physical and mental health than those without intellectual disability, including higher rates of long-term conditions and complex multi-morbidity [3, 4, 5], and die up to 20 years younger than the general population [6, 7]. This mortality gap is consistent across high-income countries [8]. Furthermore, roughly one-third of deaths of people with intellectual disability are potentially avoidable with the provision of good quality healthcare [6, 9, 10, 11]. Addressing the health inequalities experienced by this group is a priority for governments in the United Kingdom (UK) and beyond [12, 13].

People with intellectual disability are more likely to be admitted to general hospitals, where they stay longer than those without intellectual disability [14, 15]. They are at risk of receiving poor quality care and of their needs not being met for reasons that include; lack of knowledge and understanding amongst health professionals, diagnostic overshadowing, and institutional discrimination [16, 17, 18]. People with intellectual disability and their carers frequently report poor experiences of general hospital care, including inadequate communication and failure to acknowledge carers’ expertise [19, 20, 21].

Recognition of intellectual disability is essential to allow additional support needs to be identified and reasonable adjustments for these. *Healthcare for All*, a UK government-funded inquiry into access to healthcare for people with intellectual disability, highlighted the need to identify people with intellectual disability at all points of healthcare delivery, including hospital admission [17]. However, little information exists on how well intellectual disability is recognised and recorded in general hospital settings. One existing study suggests poor recording of intellectual disability in people who are admitted to hospital, but was based on ecological data which is at risk of bias [22].

In this study, we sought to:

1. investigate the recording of intellectual disability in adults with a confirmed intellectual disability diagnosis who were admitted to general hospitals
2. analyse changes in recording of intellectual disability in those admitted to hospital over time
3. identify clinical and socio-demographic factors associated with intellectual disability being unrecorded in those with the condition

## Methods

### Study setting

This study was conducted in England where most healthcare is provided by the National Health Service (NHS), a state-funded provider. Secondary (specialist) healthcare is delivered by organisations known as Trusts. Acute Trusts provide general hospital services for physical health problems, including in-patient and out-patient facilities, emergency departments, and surgical care. Mental health Trusts provide secondary mental healthcare including services for the assessment, diagnosis, and psychiatric management of people with intellectual disability.

The South London and Maudsley (SLaM) NHS Trust is one of the largest providers of secondary mental healthcare in Europe, serving a population of approximately 1.2 million people distributed between four demographically diverse south London boroughs. SLaM’s comprehensive mental healthcare services include those for diagnosis and psychiatric management of people with intellectual disability, which are provided by dedicated multi-disciplinary community learning disability teams (CLDTs) in each borough.

### Study design and data source

This is a retrospective cohort study using data from two linked clinical datasets, the SLaM Biomedical Research Centre (BRC) case register, and the English Hospital Episode Statistics (HES) database.

### South London and Maudsley Biomedical Research Centre Case Register

The SLaM BRC case register contains de-identified electronic health records (EHRs) of over 500,000 patients who have received care from any SLaM service since April 2006 [23]. Data are recorded either in structured fields (e.g. age, ethnicity, diagnosis) or as part of the unstructured free text record consisting of correspondence and other clinical case notes. The Clinical Record Interactive Search (CRIS) software was created to enable users to extract demographic and clinical information from the EHR for scientific research [23]. CRIS deploys over 100 natural language processing (NLP) algorithms developed on General Architecture for Text Engineering (GATE) software [24], developed over the last 10+ years, to extract relevant information from the free text record [25], in addition to data from structured fields.

### Hospital Episode Statistics database

Hospital Episode Statistics (HES) is a national dataset compiled by NHS general hospital providers, and curated by NHS Digital, that includes details of all admissions, out-patient appointments, and attendances to emergency departments in England [26]. HES data are used to monitor activity and as the basis for remunerating hospitals for the care they provide; they can also be used for secondary research in a fully anonymised format. We used HES inpatient admission data which include diagnoses identified during the hospital contact, recorded using codes of the International Statistical Classification of Diseases and Related Health Problems, 10^th^ edition (ICD-10) [27]. Up to 20 diagnostic codes can be added for each patient’s HES record. We also obtained admission and discharge dates, and admission route (elective/planned or non-elective/emergency).

### Study participants

We retrieved records of all adults (≥ 18 years) in the SLaM BRC case register with a diagnosis of intellectual disability who had received care from SLaM NHS Trust between 2006 and 2019. A diagnosis of intellectual disability and was taken as either a record of an ICD-10 code within the mental retardation subchapter (ICD-10 code F70 to F79) or a diagnosis of intellectual disability in the free text, extracted using the relevant NLP algorithm. These individuals’ patient records were linked with HES data over the same period using approved secure processes via the SLaM Clinical Data Linkage Service [28], to identify all general hospital admissions during the study period and diagnoses recorded within the general hospital setting during each admission.

### Co-variates

The following data were extracted from the structured fields of the SLaM BRC case register for each participant using the recording closest to their first general hospital admission: age, sex, ethnicity (white, Asian, Black, mixed, other), marital status (married, civil partnership, co-habiting, single, divorced, separated, widowed). Degree of intellectual disability (mild, moderate, severe, profound) was extracted first from the patient’s latest ICD-10 diagnostic code (where the second character denotes level of disability) or, if this was not specified, using the Health of the Nation Outcome Scales for People with Learning Disabilities (HONOS-LD), an outcome measure recommended for routine clinical use with this population [29], or from free text records. Socio-economic status of participants was estimated using the Index of Multiple Deprivation (IMD), a widely used neighbourhood-level (each area comprising approximately 1500 individuals) measure of relative deprivation based on 37 indicators related to the patient’s address [30].

### Analysis

The BRC Case Register record of intellectual disability was used as the ‘gold standard’ diagnosis against which recording of intellectual disability in general hospitals was tested. The intellectual disability services within SLaM that contribute to the BRC register specialise in diagnosis and management of people with intellectual disability. It is standard procedure to add these diagnoses to the record using a structured electronic form. Diagnosis of intellectual disability is by trained professionals who are experienced in using standard classification systems and is expected to be made following a combination of formal cognitive testing, assessment of adaptive functioning, and evidence that deficits have been present since at least childhood, so we judged that the specialist secondary mental health service would be an appropriate gold-standard.

Summary statistics were used to describe the sample. We calculated sensitivity of recording of intellectual disability in general hospital records (HES) after the first recorded diagnosis in the BRC Case Register:

a. for each admission (proportion of all admissions of people with intellectual disability during which intellectual disability is recorded) (admission sensitivity)
b. for each patient (proportion of people with intellectual disability who have the diagnosis recorded in any hospital admission) (patient-level sensitivity)
c. for emergency admissions only (proportion of all emergency (non-elective) admissions of people with intellectual disability in which intellectual disability is recorded), as the majority of elective admissions are only brief and recurrent admissions e.g. for renal dialysis or wound dressing during which we judged full diagnostic assessment may not be appropriate or customary.

For each of these we investigated intellectual disability recording in the HES data using:

i. ICD-10 intellectual disability codes (F70-F79)
ii. ICD-10 intellectual disability codes (F70-F79) and a single additional code, F81.9 (developmental disorder of scholastic skills, unspecified), which we noted during initial exploration of the data was used in a significant proportion of people with a BRC case register diagnosis

We investigated time trends in recording of intellectual disability in general hospitals by reporting the proportion of each individual’s first emergency hospital admission between 2006 and 2019 in which intellectual disability was recorded. We reported on the trend using chi-squared test for trend.

We used logistic regression to identify factors associated with intellectual disability (F70-F79) being unrecorded in HES data. Univariate regression was conducted for each variable followed by multivariable analysis adjusted for each co-variate and number of hospital admissions in the study period. Marital status was collapsed into a binary variable for this stage of the analysis (married or unmarried) in response to small numbers in some response categories. In a sensitivity analysis, we used multiple imputation by chained equations to impute missing values for all variables that contained missing data [31] and conducted logistic regression on each of twenty imputed datasets, combining coefficients using Rubin’s rules [32]. All analysis was performed using STATA v14.

### Ethics

The SLaM BRC Case Register and CRIS have received ethical approval from the Oxfordshire Research Ethics Committee C (18/SC/0372) for secondary analysis of deidentified health data. Researchers did not have access to patient-identifiable information.

## Results

### Sample characteristics

The sample comprised 2,477 adults with intellectual disability who were admitted a general hospital in England at least once over the course of the study period. Details of the sample are provided in table 1. There was a slight preponderance of males (53.9%) and the majority had mild intellectual disability (59.1% with data available). The average age at first general hospital admission was 44 years. The largest ethnic group was white. Most (83.5%) were unmarried.

**Table 1.** Demographics of adults with diagnosed intellectual disability admitted to an English general hospital during the study period (*n*=2,477)

|  |  | n | % |
| --- | --- | --- | --- |
| **Age** | Mean (SD) | 44.0 (16.1) | - |
| **Degree of intellectual disability** | Mild | 928 | 37.5 |
|  | Moderate | 420 | 17.0 |
|  | Severe | 208 | 8.4 |
|  | Profound | 13 | 0.5 |
|  | Missing | 908 | 36.7 |
| **Sex** | Male | 1,335 | 53.9 |
|  | Female | 1,142 | 46.1 |
| **Ethnicity** | White | 1,517 | 61.2 |
|  | Asian | 114 | 4.6 |
|  | Black | 539 | 21.8 |
|  | Mixed | 73 | 3.0 |
|  | Other | 56 | 2.3 |
|  | Missing | 178 | 7.2 |
| **Marital status\*** | Married | 112 | 4.5 |
|  | Unmarried | 2,068 | 83.5 |
|  | Missing | 297 | 12.0 |
| **Deprivation score\*\*** | Mean (SD) | 29.1 (10.7) | - |
|  | Missing | 71 | - |
| **Number of admissions** | Range | 1-740 | - |
|  | Median (IQR) | 5 (3-10) | - |

\*married includes civil partner, co-habiting; unmarried includes single, widowed, divorced
\*\*deprivation score is the Index of Multiple Deprivation. Higher scores indicate a greater degree of deprivation.

There were 27,314 discrete admissions to general hospitals over the study period; 16,270 were non-elective admissions and the remainder (11,044) were elective admissions (e.g. for planned surgery, routine dialysis, or change of wound dressing). The median number of total admissions per patient was 5.

### Sensitivity of general hospital recording of intellectual disability

Taking each of the 27,314 admissions independently, 788 had a HES record of intellectual disability (F70-79) (admission-level sensitivity = 2.9%, 95%CI = 2.7, 3.1). Of each emergency admission (*n*= 16,270), 319 had a record of intellectual disability (sensitivity = 2.0%, 95%CI 1.8, 2.2). Of the 2,477 people who were admitted to hospital, 445 had a record of intellectual disability defined using intellectual disability codes (F70-79) at any time in their general hospital record (patient-level sensitivity = 18.0%, 95%CI = 16.5, 19.5).

Including the ICD-10 code F81.9 (developmental disorder of scholastic skills, unspecified) in addition to the F70-79 codes resulted in a notable increase in those with intellectual disability who were recorded in the general hospital record (admission-level sensitivity = 27.7% (27.2, 28.3); patient-level sensitivity = 66.3% (64.4, 68.1) (table 2).

**Table 2.** Sensitivity of recording of intellectual disability in English general hospital records 2006-2019 for individual admissions and for individual patients by ICD-10 code group

|  | Sensitivity (95% CI) |  |
| --- | --- | --- |
| ICD-10 descriptor | Intellectual disability | Intellectual disability<br>Developmental disorder of scholastic skills, unspecified |
| ICD-10 codes | F70-79 | F70-79 and F81.9 |
| For each admission<br>(admission sensitivity) | 2.9 (2.7, 3.1) | 27.7 (27.2, 28.3) |
| For each patient<br>(patient-level sensitivity) | 18.0 (16.5, 19.5) | 66.3 (64.4, 68.1) |
| For each admission<br>(emergency admissions only) | 2.0 (1.8, 2.2) | 32.7 (32.0, 33.4) |

### Time trends in recording intellectual disability in general hospital records

Data for recording of intellectual disability stratified by year for the first emergency admission of each patient are shown in Fig 1. Strict recording of intellectual disability using F70-79 codes showed little overall change over the study period. Including the F81.9 code with F70-F79 codes showed a consistent increase in those who were identified, from 17.5% (95% CI = 13.8, 21.7) in 2005 to 62.5% (40.6, 81.2) in 2019 (χ^2^ for trend = 138.7, p <0.0001). Raw data and proportions are given in supporting information (table S1).

**Fig 1.**
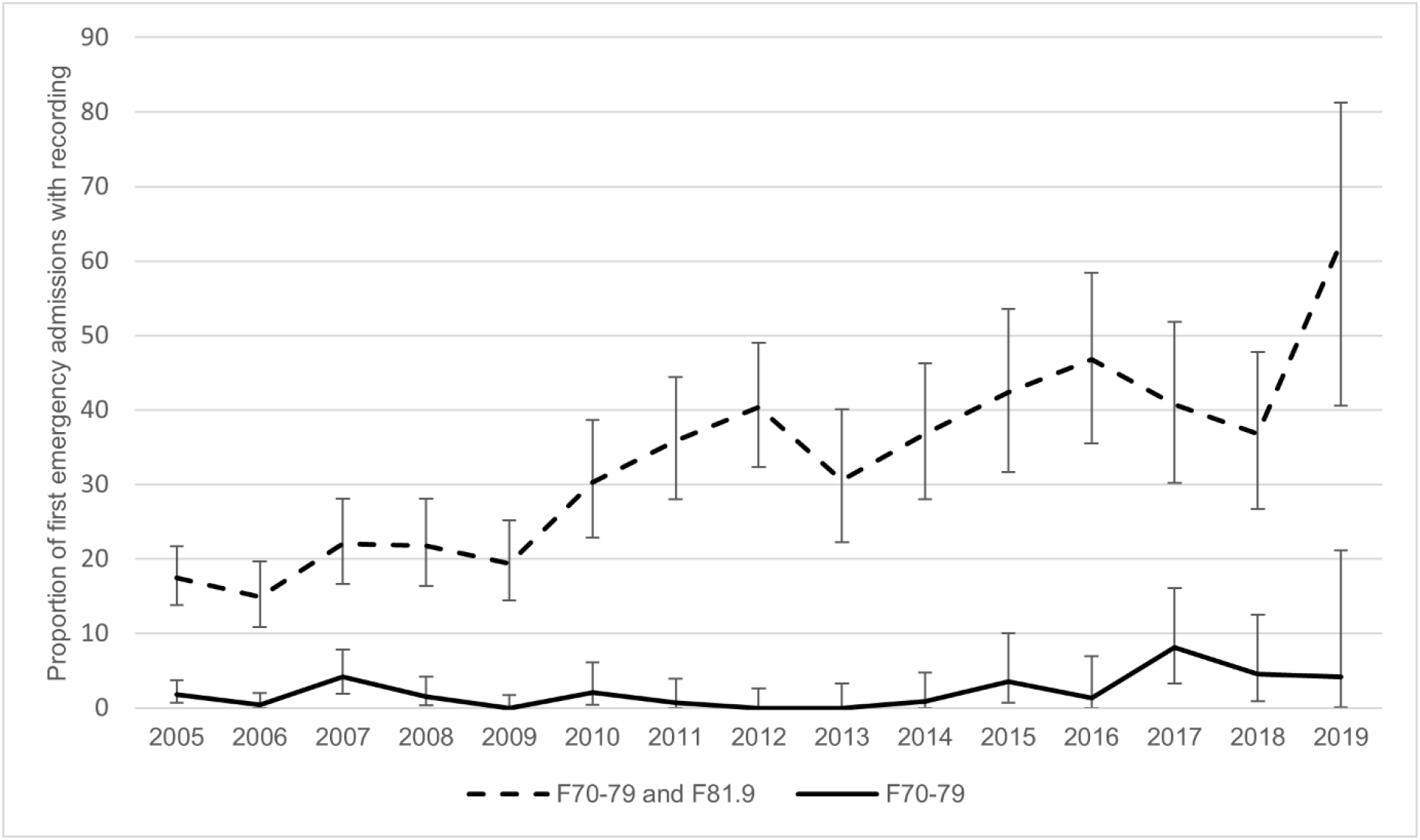
Time trends in recording of intellectual disability in those admitted to a general hospital in England, 2005-2019

### Associations with intellectual disability being unrecorded in hospital records

Factors associated with a person with intellectual disability never having this accurately recorded (as ICD-10 codes F70-F79) in their general hospital record were investigated (table 3). In the adjusted analysis, having more severe intellectual disability was associated with lower odds of intellectual disability being unrecorded (adjusted odds ratio (aOR) for severe vs mild intellectual disability 0.30 (0.20, 0.46), *p*<0.001), and being married was associated with higher odds of intellectual disability being unrecorded (aOR 3.12, 95%CI 1.10, 8.81, *p*=0.03). Regression results with imputed values were similar and are given in supporting information (table S2).

**Table 3.**
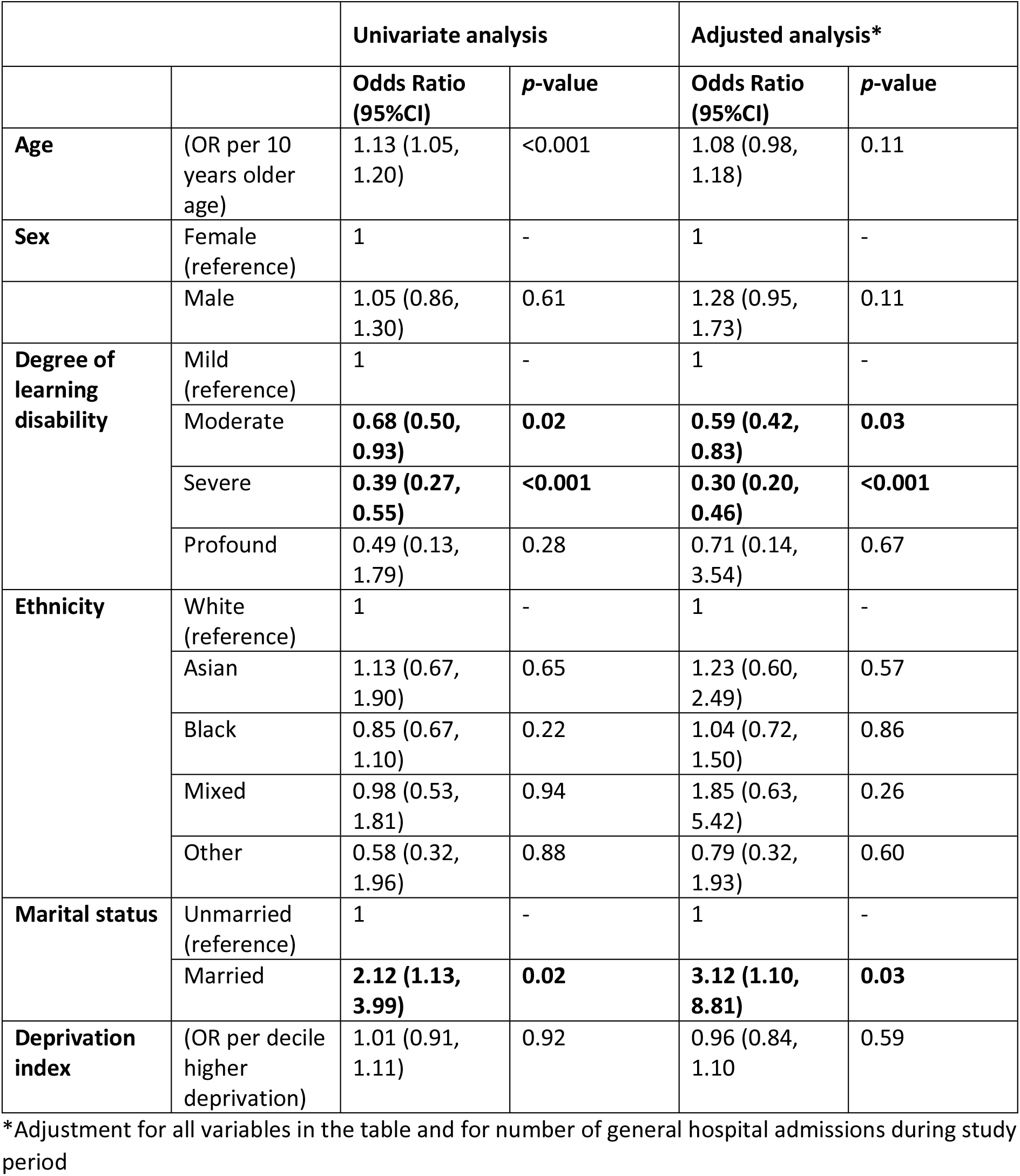
Odds of intellectual disability being unrecorded in the general hospital record of adults with intellectual disability attending hospital

|  |  | Univariate analysis |  | Adjusted analysis* |  |
| --- | --- | --- | --- | --- | --- |
|  |  | Odds Ratio<br>(95%CI) | p-value | Odds Ratio<br>(95%CI) | p-value |
| **Age** | (OR per 10 years older age) | 1.13 (1.05, 1.20) | <0.001 | 1.08 (0.98, 1.18) | 0.11 |
| **Sex** | Female (reference) | 1 | - | 1 | - |
|  | Male | 1.05 (0.86, 1.30) | 0.61 | 1.28 (0.95, 1.73) | 0.11 |
| **Degree of learning disability** | Mild (reference) | 1 | - | 1 | - |
|  | Moderate | **0.68 (0.50, 0.93)** | **0.02** | **0.59 (0.42, 0.83)** | **0.03** |
|  | Severe | **0.39 (0.27, 0.55)** | **<0.001** | **0.30 (0.20, 0.46)** | **<0.001** |
|  | Profound | 0.49 (0.13, 1.79) | 0.28 | 0.71 (0.14, 3.54) | 0.67 |
| **Ethnicity** | White (reference) | 1 | - | 1 | - |
|  | Asian | 1.13 (0.67, 1.90) | 0.65 | 1.23 (0.60, 2.49) | 0.57 |
|  | Black | 0.85 (0.67, 1.10) | 0.22 | 1.04 (0.72, 1.50) | 0.86 |
|  | Mixed | 0.98 (0.53, 1.81) | 0.94 | 1.85 (0.63, 5.42) | 0.26 |
|  | Other | 0.58 (0.32, 1.96) | 0.88 | 0.79 (0.32, 1.93) | 0.60 |
| **Marital status** | Unmarried (reference) | 1 | - | 1 | - |
|  | Married | **2.12 (1.13, 3.99)** | **0.02** | **3.12 (1.10, 8.81)** | **0.03** |
| **Deprivation index** | (OR per decile higher deprivation) | 1.01 (0.91, 1.11) | 0.92 | 0.96 (0.84, 1.10) | 0.59 |

\*Adjustment for all variables in the table and for number of general hospital admissions during study period

## Discussion

Accurate recognition and recording of intellectual disability in people who are admitted to general hospitals is important so that additional support needs can be identified and the necessary adaptations to care and processes can be provided. These might include communication support, use of the Mental Capacity Act [33], involving family or paid carers, and addressing issues around planning safe discharge [34]. Collecting and recording accurate data is also important on a population level to allow healthcare providers and commissioners to understand their patient group, allocate adequate resource, and plan services [17].

Using large, linked datasets of real-world clinical data, we found that intellectual disability was poorly recorded in people who were admitted to general hospitals. Taking each admission individually, intellectual disability was accurately recorded in under 3% of all general hospital admissions and just under one-fifth of those with a confirmed diagnosis of intellectual disability (as recorded by specialist intellectual disability and mental health services) ever had the condition accurately recorded in their general hospital records, despite them having several admissions on average. These findings may reflect poor knowledge and recognition of intellectual disability amongst general hospital staff, a reluctance to label patients with disabilities, or a lack of understanding of why this is necessary.

Low rates of recording may also reflect clinical coding errors. We observed a proportion of adults with intellectual disability who were coded as having learning *difficulty* using the ICD-10 descriptor ‘developmental disorder of scholastic skills, unspecified’. This diagnostic category is ill-defined, and use is discouraged [27] but it might be applied when there are specific deficits in language and speech development, motor co-ordination, or the development of arithmetic, reading, or spelling proficiency without global intellectual impairment. Regular use of this code suggests that some form of cognitive impairment was more frequently recognised during the admission; in practice, this code seems to be used as a proxy for intellectual disability. However, even including the additional code, one third of people with intellectual disability never had a relevant diagnosis recorded, and these diagnoses were unrecorded in around three-quarters of admissions. These codes are also likely to be missed in national administrative returns, leading to under-reporting of healthcare use by people with intellectual disability. From a research perspective, using a strict definition of intellectual disability (F70-F79) in future studies using HES will miss a substantial proportion of people who likely should be included.

There is no benchmark against which the results of this study can be directly compared, but our figures show that recording of intellectual disability within general hospitals is lower than recording of other mental disorders. Similar methodology has reported recording rates of 78% for dementia and 56% for schizophrenia [36]. Missed recording of intellectual disability is also not only a problem in secondary care; far fewer people than would be expected are included on intellectual disability registers held by primary care in England [37].

Our data indicate that recording of intellectual disability has improved over time only if the related (but technically incorrect) code of learning difficulty (ICD-10 F81.9) is included. Several factors may have contributed to this increased recording, including the presence of learning disability liaison nurses in general hospitals and generally increased awareness of the health needs of people with intellectual disability [38, 39]. This trend may also reflect overall improvements in diagnostic coding in HES data that has been observed over time [40].

Analysis of factors associated with unrecorded diagnosis showed that having a less severe intellectual disability was independently associated with increased likelihood of intellectual disability never being recorded in general hospital data. Mild intellectual disability may not be immediately obvious on meeting a person with the condition and specific enquiry may not be included in standard medical admission assessments. People with intellectual disability who were married, cohabiting, or in a civil partnership were also more likely *not* to have their intellectual disability recorded; it may be that assumptions about the lives of people with intellectual disability contributed to this finding, or that these people had milder intellectual disability which was more easily missed, even though we adjusted for intellectual disability severity.

Recording of intellectual disability was similar in non-elective (emergency) admissions as in elective (planned) admissions. Other studies have shown the recording of dementia and severe mental illness to be better in non-elective admissions [35, 36]; the universally low rate of recording of intellectual disability demonstrates further awareness of the condition in general hospitals is needed.

### Strengths and limitations

In this study we were able to identify a large representative cohort using a local specialist mental health and community intellectual disability team case register and link this to a hospital admissions database with national coverage. The results add to the very limited existing data on the recognition and recording of intellectual disability in general hospitals in England and will provide impetus for improvements.

Our study has some limitations. We used diagnosis of intellectual disability made by a secondary mental health care service (which includes specialist intellectual disability teams) as the gold-standard meaning that only individuals with intellectual disability living in the catchment area who have accessed secondary mental health and intellectual disability services have been included.

However, our use of specialist service data, interrogation of structured fields and free-text records using natural language processing improves confidence in the diagnosis and this approach has been validated in other mental disorders showing high precision [28]. Our approach allowed the construction of a large cohort representative of people with clinically diagnosed intellectual disability, which would not have been possible had we assessed all participants with a standardised assessment.

It is possible that some people with intellectual disability, particularly those with specific genetic conditions such as Down syndrome, were accurately recorded with codes outside the generic ICD-10 intellectual disability codes of F70-F79. This may have caused an under-estimate of the sensitivity of hospital recording, although only a minority (roughly one-quarter) of people with intellectual disability have a recognised genetic cause that is eligible to be coded separately, so this would not account for the degree of under-recording we observed. Furthermore, not all people with genetic conditions associated with intellectual disability do in fact have intellectual disability, therefore a code for intellectual disability should still be used where it is relevant.

This study looks only at recording of intellectual disability in hospital records, using the patient discharge summary as the source of HES data. Whilst helpful as a first step in contemplating and instituting reasonable adjustments, recording does not guarantee that adapted person-centred care will be provided, and we have no measure of the quality of care that was provided. Similarly, it is possible that in some cases where intellectual disability was not recorded, this was due to a failure of the administrative process alone, and that intellectual disability was indeed recognised and managed appropriately during the admission. Even in these cases, however, it is still necessary from a service-level and surveillance perspective that intellectual disability is documented.

### Clinical and research implications

There is a need for improved recognition and recording of intellectual disability in general hospitals to improve hospital outcomes and care experiences. An active approach to identification has been suggested, with routine screening questions being asked at entry points to care [41]. Pre-admission assessments and learning disability identification checklists are recommended to ensure that people with intellectual disability receive the support they require in hospital and to give advance notice to staff [42, 43] but are only possible in the case of planned admissions. People with intellectual disability are more likely than the general population to present to emergency care [44, 45]; automatic flagging of people with intellectual disability attending hospital could be achieved with better integration of health records between statutory services, underpinned by stringent data sharing protocols.

It is important that recognition of intellectual disability is supplemented by staff understanding of the range of health and communication needs that people with intellectual disability may have. Interventions that have been shown to be effective then need to be implemented, such as involvement of learning disability liaison nurses and adapted communication approaches. A programme of mandatory training in autism and learning disability for all staff will soon be introduced across the UK National Health Service [46] and is hoped will contribute to improvements in care.

## Conclusions

This study demonstrates that, currently, HES data alone cannot be used to identify a cohort of people with intellectual disability attending hospital, as a significant proportion would be missed. Using a range of sources with health database linkage can increase coverage and provide a powerful tool for epidemiological research to drive improvements in care in this group.

Future studies could investigate the experience and outcomes of care between those who were recorded as having intellectual disability during their admission and those who are not. Similar work could investigate recording of intellectual disability in out-patient clinics. Hospital recognition of other developmental disorders in whom health inequalities also exist, such as autism and borderline intellectual functioning, could also be investigated [47, 48].

## Data Availability

The data used in this work have been obtained from the Clinical Record Interactive Search (CRIS), a system that has been developed for use within the NIHR Mental Health Biomedical Research Centre (BRC) at the South London and Maudsley NHS Foundation Trust (SLaM), and Hospital Episode Statistics (HES), a dataset collated by NHS Digital. Individual-level data are restricted in accordance with patient led governance established at SLaM, and by NHS Digital in the case of linked HES data. CRIS data are available for authorised researchers who meet the criteria for access as (1) SLaM employees or (2) those having an honorary contract or letter of access from SLaM. Further details can be obtained by e-mailing

## Supporting information

**S1 Table.** Time trends in recording intellectual disability in adults admitted to English general hospitals, 2005-2019

|  | 2005 | 2006 | 2007 | 2008 | 2009 | 2010 | 2011 | 2012 | 2013 | 2014 | 2015 | 2016 | 2017 | 2018 | 2019 | Total |
| --- | --- | --- | --- | --- | --- | --- | --- | --- | --- | --- | --- | --- | --- | --- | --- | --- |
| **F70-F79** |  |  |  |  |  |  |  |  |  |  |  |  |  |  |  |  |
| No (FN) | 375 | 274 | 205 | 203 | 222 | 139 | 141 | 141 | 111 | 116 | 82 | 78 | 79 | 64 | 23 | 2253 |
| Yes (TP) | 7 | 1 | 9 | 3 | 0 | 3 | 1 | 0 | 0 | 1 | 3 | 1 | 7 | 3 | 1 | 40 |
| Total | 382 | 275 | 214 | 206 | 222 | 142 | 142 | 141 | 111 | 117 | 85 | 79 | 86 | 67 | 24 | 2293 |
| **F70-79 and F81** |  |  |  |  |  |  |  |  |  |  |  |  |  |  |  |  |
| No (FN) | 315 | 234 | 167 | 161 | 179 | 99 | 91 | 84 | 77 | 74 | 49 | 42 | 51 | 35 | 9 | 1667 |
| Yes (TP) | 67 | 41 | 47 | 45 | 43 | 43 | 51 | 57 | 34 | 43 | 36 | 37 | 35 | 32 | 15 | 626 |
| Total | 382 | 275 | 214 | 206 | 222 | 142 | 142 | 141 | 111 | 117 | 85 | 79 | 86 | 67 | 24 | 2293 |

FN, false negative; TP, true positive

|  | 2005 | 2006 | 2007 | 2008 | 2009 | 2010 | 2011 | 2012 | 2013 | 2014 | 2015 | 2016 | 2017 | 2018 | 2019 |
| --- | --- | --- | --- | --- | --- | --- | --- | --- | --- | --- | --- | --- | --- | --- | --- |
| **F70-F79** |  |  |  |  |  |  |  |  |  |  |  |  |  |  |  |
| Proportion recorded (95%CI) | 1.8<br>(0.7, 3.7) | 0.4<br>(0.0, 2.0) | 4.2<br>(1.9, 7.8) | 1.5<br>(0.3, 4.2) | 0.0<br>(0.0, 1.7) | 2.1<br>(0.4, 6.1) | 0.7<br>(0.0, 3.9) | 0.0<br>(0.0, 2.6) | 0.0<br>(0.0, 3.3) | 0.9<br>(0.0, 4.7) | 3.5<br>(0.7, 10.0) | 1.3<br>(0.0, 6.9) | 8.1<br>(3.3, 16.1) | 4.5<br>(0.9, 12.5) | 4.2<br>(0.1, 21.1) |
| **F70-79 and F81** |  |  |  |  |  |  |  |  |  |  |  |  |  |  |  |
| Proportion recorded (95%CI) | 17.5<br>(13.8, 21.7) | 14.9<br>(10.9, 19.7) | 22.0<br>(16.6, 28.1) | 21.8<br>(16.4, 28.1) | 19.4<br>(14.4, 25.2) | 30.3<br>(22.9, 38.6) | 35.9<br>(28.0, 44.4) | 40.4<br>(32.3, 49.0) | 30.6<br>(22.2, 40.1) | 36.8<br>(28.0, 46.2) | 42.4<br>(31.7, 53.6) | 46.8<br>(35.5, 58.4) | 40.7<br>(30.2, 51.8) | 36.8<br>(26.7, 47.8) | 62.5<br>(40.6, 81.2) |

CI, confidence interval

**S2 Table.**
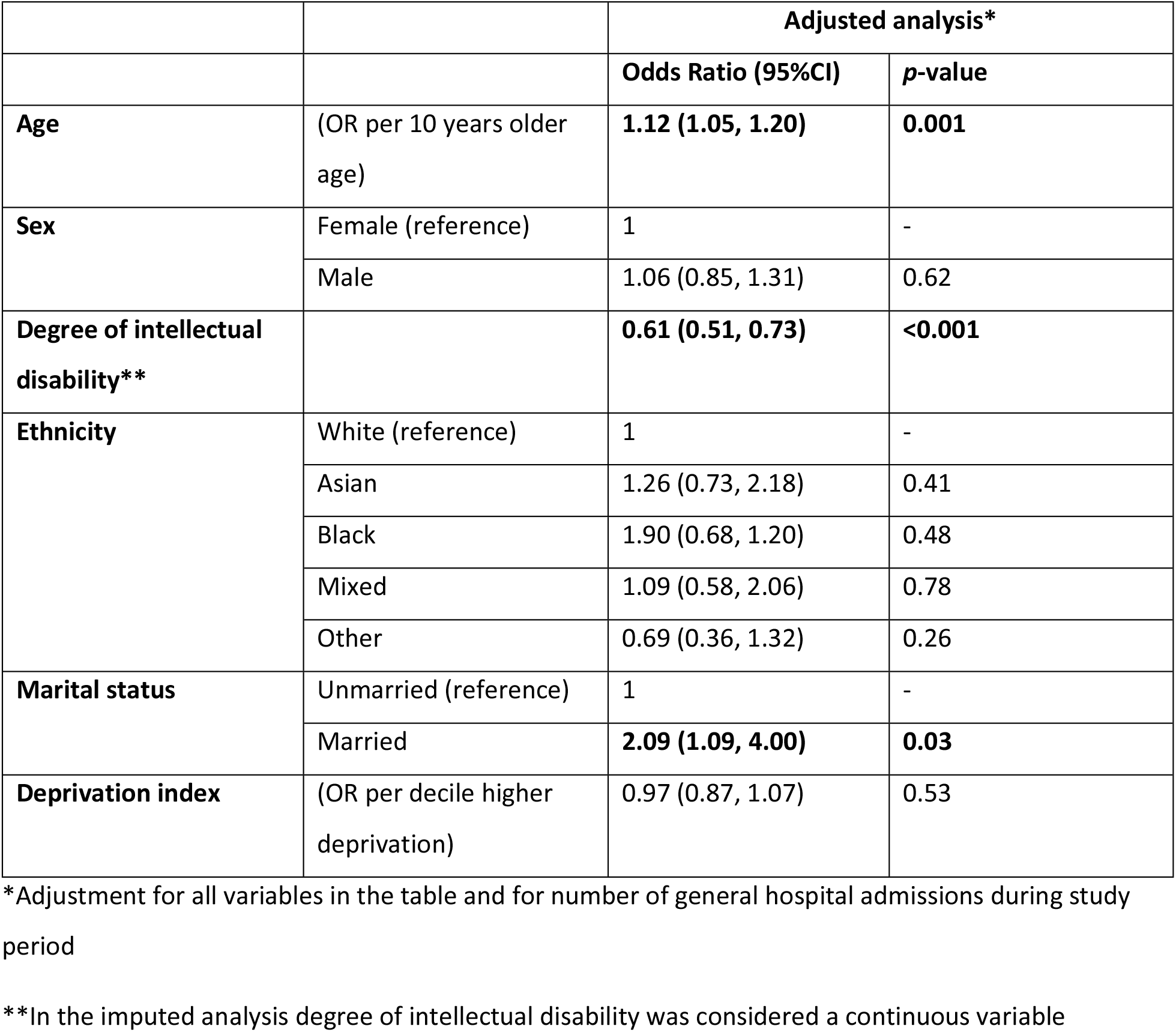
Odds of intellectual disability being unrecorded in the general hospital record of adults with intellectual disability attending hospital (with multiple imputation for missing data)

|  |  | **Adjusted analysis\*** |  |
| --- | --- | --- | --- |
|  |  | **Odds Ratio (95%CI)** | **p-value** |
| **Age** | (OR per 10 years older age) | **1.12 (1.05, 1.20)** | **0.001** |
| **Sex** | Female (reference) | 1 | - |
|  | Male | 1.06 (0.85, 1.31) | 0.62 |
| **Degree of intellectual disability\*\*** |  | **0.61 (0.51, 0.73)** | **<0.001** |
| **Ethnicity** | White (reference) | 1 | - |
|  | Asian | 1.26 (0.73, 2.18) | 0.41 |
|  | Black | 1.90 (0.68, 1.20) | 0.48 |
|  | Mixed | 1.09 (0.58, 2.06) | 0.78 |
|  | Other | 0.69 (0.36, 1.32) | 0.26 |
| **Marital status** | Unmarried (reference) | 1 | - |
|  | Married | **2.09 (1.09, 4.00)** | **0.03** |
| **Deprivation index** | (OR per decile higher deprivation) | 0.97 (0.87, 1.07) | 0.53 |

\*Adjustment for all variables in the table and for number of general hospital admissions during study period
\*\*In the imputed analysis degree of intellectual disability was considered a continuous variable

## Notes

### Competing Interest Statement

The authors have declared no competing interest.

### Funding Statement

The authors received no specific funding for this work. RSt is part-funded by: i) the National Institute for Health Research (NIHR) Biomedical Research Centre at the South London and Maudsley NHS Foundation Trust and King’s College London ii) the National Institute for Health Research (NIHR) Applied Research Collaboration South London (NIHR ARC South London) at King’s College Hospital NHS Foundation Trust iii) the DATAMIND HDR UK Mental Health Data Hub (MRC grant MR/W014386).

